# Testing for coronavirus (SARS-CoV-2) infection in populations with low infection prevalence: the largely ignored problem of false positives and the value of repeat testing

**DOI:** 10.1101/2020.08.19.20178137

**Authors:** Cathie Sudlow, Peter Diggle, Oliver Warlow, David Seymour, Ben Gordon, Rhos Walker, Charles Warlow

**Author notes:** Correspondence to: Prof Cathie Sudlow, BHF Data Science Centre, Health Data Research UK.

## Abstract

**Background:** Calls are increasing for widespread SARS-CoV-2 infection testing of people from populations with a very low prevalence of infection. We quantified the impact of less than perfect diagnostic test accuracy on populations, and on individuals, in low prevalence settings, focusing on false positives and the role of confirmatory testing.

**Methods:** We developed a simple, interactive tool to assess the impact of different combinations of test sensitivity, specificity and infection prevalence in a notional population of 100,000. We derived numbers of true positives, true negatives, false positives and false negatives, positive predictive value (PPV – the percentage of test positives that are true positives) and overall test accuracy for three testing strategies: (1) single test for all; (2) add repeat testing in test positives; (3) add further repeat testing in those with discrepant results. We also assessed the impact on test results for individuals having one, two or three tests under these three strategies.

**Results:** With sensitivity of 80%, infection prevalence of 1 in 2,000, and specificity 99.9% on all tests, PPV in the tested population of 100,000 will be only 29% with one test, increasing to > 99.5% (100% when rounded to the nearest %) with repeat testing in strategies 2 or 3. More realistically, if specificity is 95% for the first and 99.9% for subsequent tests, single test PPV will be only 1%, increasing to 86% with repeat testing in strategy 2, or 79% with strategy 3 (albeit with 6 fewer false negatives than strategy 2). In the whole population, or in particular individuals, PPV increases as infection becomes more common in the population but falls to unacceptably low levels with lower test specificity.

**Conclusion:** To avoid multiple unnecessary restrictions on whole populations, and in particular individuals, from widespread population testing for SARS-CoV-2, the crucial roles of extremely high test specificity and of confirmatory testing must be fully appreciated and incorporated into policy decisions.

## BACKGROUND

### No diagnostic test is perfect

However accurately some diagnostic tests may perform in well-controlled laboratory conditions, no test is 100% accurate in real-world clinical settings. Most science and media coverage and debate about the accuracy of molecular testing for SARS-CoV-2 coronavirus infection has focused on the problem of ‘false negatives’, where people with the virus have a negative test result when they should test positive. A perfect test would be 100% sensitive – i.e., all cases infected with the virus should test positive. There are several reasons for less than 100% sensitivity with molecular tests for current SARS-CoV-2 infection (as distinct from antibody tests, aimed at detecting evidence of prior infection), including imperfect nasal and throat swabbing technique, insufficient virus present, laboratory error, and timing (i.e., after infection, the virus may take some time to appear in the nose and throat and then disappears at some point during the course of the infection).

One unfortunate consequence of false negatives is that people who really are infected with the virus may be reassured by a negative test (especially if they are asymptomatic or pre-symptomatic) and go on to infect others, perhaps by going back to work in a care home or factory where social distancing is challenging. However, the false negative problem is far less of an issue if someone is very likely to have SARS-CoV-2 coronavirus infection, for example if he or she is in a care home with many other residents testing positive, and has a cough, breathlessness, fever and hypoxia. The negative test would not put a doctor off diagnosing SARS-CoV-2 coronavirus infection (“if it looks like a duck and quacks it probably is a duck”), managing the patient appropriately, and maybe ordering another test. In other words, experienced clinicians will generally (and quite appropriately) ignore a negative test if on clinical grounds infection is highly likely.

### False positive test results are common when infection is uncommon

By contrast, there has been much less discussion about the problem of ‘false positives’, which relate to what is referred to as the *specificity* of the test. A 100% specific test, the perfect test, would be negative in everyone tested who did *not* have the virus. Anything less than 100% specificity inevitably leads to some truly virus-free people testing positive: false positives. Possible reasons for a false positive result include cross reactivity with another virus, sample contamination, and laboratory error.

Unfortunately, as the true frequency (or prevalence) of infection becomes less and less in any population, the problem of false positive tests becomes far more of an issue, even with a highly specific test. The lower the prevalence, the higher will be the proportion of false positive results among all the positive results of even an almost perfectly specific test. This relationship is incredibly important for countries such as the UK (and many others across Europe and elsewhere), where the percentage of individuals in the community who are actively infected is currently very low (< 0.1% in the UK in late July),^1^ but where increasing numbers of prominent scientists and politicians are calling for widespread testing of asymptomatic people from these low prevalence populations.^2–6^

We aimed to provide a simple, transparent tool to help policy makers, the scientific community, health professionals, journalists and the public understand the problem of incorrect test results, and to demonstrate how this problem may be addressed by conducting a second or even third test. In particular, we wished to highlight the issue of false positive results in low prevalence settings, and to draw attention to the possible unintended adverse consequences for populations and individuals if these are not fully understood.

## METHODS

We implemented a probabilistic calculation as an interactive tool within a Microsoft Excel spreadsheet workbook (https://www.hdruk.ac.uk/projects/false-positives/). The tool requires three input parameters: the prevalence of infection in the population from which the individuals being tested are drawn; test sensitivity; and test specificity. These parameters can be varied interactively so that the results of different scenarios can be examined. We present the results in three different formats:

i. to assess population level impact and inform population level policies, the numbers of people from a population of 100,000, all of whom are tested, who end up with true positive, true negative, false positive and false negative status after testing strategies with one, two or three rounds of testing (the ‘model mass testing’ worksheet);
ii. to assess impact on and inform guidance for individuals, the interpretation of test results for an individual, based on one, two or three tests (the ‘model individual testing’ worksheet);
iii. graphs showing the relationship between the prevalence of infection in the population and the probability of an individual with a positive test result truly having infection, where the test result is based on either one positive test only or on two positive tests (the ‘graphs’ worksheet).

For all of these output formats, repeated tests are assumed to be conducted on the same day, as the tool is designed to address the issue of incorrect test results at a particular time point, rather than to deal with variation in the probability of detecting the virus at different times during the course of infection. The repeat testing strategy is designed to be appropriate for a low prevalence situation, where all people with a positive test in the first round have a second test (in an attempt to confirm positivity and so minimize false positives), and all people with a negative test in the second round have a third test (in an attempt to resolve the situation where tests from rounds one and two yield discrepant results).

The tool can be used to illustrate a wide range of scenarios for different potential values of infection prevalence, sensitivity and specificity. Here we use the tool’s outputs to illustrate a few plausible scenarios that could arise when population prevalence of infection is low, as is currently the case in the UK.

## Choice of parameters for illustrative scenarios

### Prevalence

Since mid-May, the Office for National Statistics has published the results of weekly coronavirus infection surveys, reporting that between 1 in 4000 and 1 in 1000 people within the community population in England were infected with SARS-CoV-2 during any one of the weeks from mid-May to early August.^1^ For our initial illustrative scenario, we assume that about 1 in 2,000 people in the population (i.e., 0.05%) are infected. To illustrate what might happen with testing in a higher prevalence setting (e.g., a different country with higher prevalence of infection in the community, a town that has had a series of small outbreaks, or a large school where several pupils are thought to have been infected), we also consider scenarios where the prevalence is 20 times higher (i.e., 1% or 1 in 100). For graphical outputs, we consider prevalence values ranging from 0.05–10%.

### Test specificity

Independent evaluations of molecular assays for SARS-CoV-2 infection performed in a single university hospital laboratory have found that around 95 to 100 % of samples with no virus present tested negative with the various molecular assays for SARS-CoV-2 infection.^7^ For our initial illustrative scenario we assume that the best test specificity possible is 99.9% and show the highly optimistic results that would arise with a test specificity of 99.9% for all three rounds of testing. A specificity of 99.9% means that 0.1% (100 – 99.9%) of all truly virus-free people tested will have a falsely positive test, i.e., one false positive result out of every 1000 virus-free people tested. We also show results for a more plausible specificity of 95% for the first round of tests and 99.9% for the smaller number of second and third round tests, assuming that these are conducted with highly optimized sample collection, sample transfer and laboratory analysis procedures. Finally, we show results arising when we consider prevalence values ranging from 0.05–10%. specificity is 95% for all three rounds of testing. For graphical outputs, we consider specificities in the range 90–99.9%.

### Test sensitivity

The same independent evaluation source reported that around 90 to 100 % of samples with the virus present tested positive,^7^ although others have reported test sensitivity to be highly variable and often lower in clinical practice (from around 30 to 90%).^6^ We show here the results for a sensitivity level of 80%, based on reports of test performance in clinical practice from the more promising end of the spectrum.

## RESULTS

For six scenarios, based on six different combinations of parameters, Table 1 shows, for each testing strategy applied to our population of 100,000, the number of tests performed, the expected numbers of people with true positive, true negative, false positive and false negative results, the percentage of all positive results that are true positives (positive predictive value [PPV]), and the overall testing accuracy (the percentage of all test results that are correct).

Below, we work through the derivation of the numbers of people assigned true or false positive or negative status with the three testing strategies for our initial illustrative scenario (shown at the top left of Table 1), which assumes an infection prevalence in the population of 0.05% (i.e. 50 of the 100,000 people truly have infection and 99,950 do not), test sensitivity of 80% and specificity of 99.9% for all three rounds of tests. In what follows, all of the quoted numbers of people whose test results fall into the different categories discussed are what we would expect to happen on average. In reality, the results of testing a finite number of people are inevitably subject to random variation.

## Strategy 1: testing everyone in the population once

After testing all 100,000 people, the expected outcome is that:

- the number of people with a true positive result will be 80% (the sensitivity) of the 50 people with infection, i.e. 40;
- this will leave 10 people who do actually have infection with a false negative test;
- the number of people with a true negative result will be 99.9 % (the specificity) of the 99,950 without infection, i.e. 99,850;
- this will leave 100 people who do not have infection but with a false positive test.

Hence 140 people will have a positive test, of whom 40 (29% of the positive tests) will be true positives and 100 (71%) will be false positives. On the other hand, 99,860 people will have a negative test, of whom 99,850 will be true negatives and only 10 will be false negatives (i.e., missed cases of infection).

## Strategy 2 – add immediate re-testing of all those with a *positive* result on their first test

Since such a large percentage of the 140 positive results of the first round of testing are false positives, a sensible strategy is to conduct a further test (immediately) on these 140 people. In this illustrative scenario, test sensitivity and specificity remain the same, but now the prevalence of infection in the tested population is much higher, 40 in 140 (29%). After re-testing these 140 people, the expected outcome is that:

- the number of people with a second true positive result will be 80% (the sensitivity) of the 40 people with infection, i.e. 32;
- this will leave 8 people who do actually have infection with a false negative test;
- the number of people with a true negative results will be 99.9% (the specificity) of the 100 without infection, which (to the nearest whole number) is 100 who now have a true negative result;
- this will leave no people with a false positive test

Taking into account the revised results in the 140 people that have been re-tested, among all 100,000 people there are now 32 people with confirmed positive status with two positive tests (all of whom are true positives), 18 with false negative status (10 from the first round and now 8 more from the second); none with false positive status; and 99,950 with true negative status (99,850 from the first round and an additional 100 from the second round).

## Strategy 3 – add further immediate re-testing of all those with a *negative* result on their second test (i.e. a positive and then a negative result)

Although the second round of testing reduced the proportion of those whose positive result was a false positive from 71% to 0%, there may be concerns about the increase in false negatives (missed cases of infection), which in this population of 100,000 people has almost doubled after the second round of testing from 10 to 18. This concern can be addressed - to some extent, at least – by conducting a further set of tests in the 108 people whose second test was negative (i.e. those with discrepant results from the first and second rounds). Of these 108 people, 8 are true cases, so the prevalence of infection for this round of testing is around 7%. After re-testing these 108 people, the expected outcome is that:

- at 80% sensitivity (given that we are rounding to the nearest whole number of people), 6 of the 8 true cases will now be detected as true positives, leaving 2 false negatives;
- at 99.9% specificity, the remaining 100 cases without infection will all have a true negative result, and so no false positives are generated.

Taking into account the revised results in the 108 people that have been re-tested again, there are now 38 people with true positive status (32 confirmed in the second round and 6 more from this third round), 12 with false negative status (10 from the first round and 2 more from this third round), none with false positive status (unchanged from the end of the second round); and 99,950 with true negative status.

## Testing a population under different scenarios

The remainder of Table 1 shows the results of this same reasoning process applied to the other scenarios. It also shows the additional number of tests required for rounds 2 and 3 of testing to enable strategies 2 and 3, as well as the PPV (i.e. the percentage of positive results that are true positives) and the overall accuracy (the percentage of all test results that are correct) for each strategy. All scenarios yield high overall accuracy, because at low prevalence most people are correctly identified as negative. This does not helpfully distinguish the pros and cons of different testing strategies. However, the results in Table 1 also show how the PPV increases at higher population infection prevalence. The critical importance of extremely high specificity in these low prevalence settings is also demonstrated: only when specificity is 99.9%, either across all rounds of testing or - more realistically - for the smaller numbers of round two and three tests, does the PPV reach around 80% or higher. Where all rounds of testing have specificities of 95% (which is also plausible in real-world settings), the PPV may be as low as 11% at best when prevalence is 1 in 2,000 (0.05%) or 72% at best when prevalence is 1 in 100 (1%). Finally, despite needing more tests, adding the third round of testing leads to no improvement and/or a slight reduction in the PPV. The advantage of strategy 3 is a small reduction in the number of false negatives (i.e. missed cases of infection), but at the cost of a more complex strategy, more tests, and more people with incorrect positive test status (false positives).

**Table 1.** Results of different scenarios for testing 100,000 people*

|  | Infection prevalence 0.05% |  |  | Infection prevalence 1% |  |  |
| --- | --- | --- | --- | --- | --- | --- |
|  | Strategy 1<sup>†</sup> | Strategy 2<sup>‡</sup> | Strategy 3 | Strategy 1<sup>†</sup> | Strategy 2<sup>‡</sup> | Strategy 3 |
| **Specificity 99.9% for all tests** |  |  |  |  |  |  |
| Repeat tests added (n)<sup>¶</sup> | 0 | 140 | 108 | 0 | 899 | 259 |
| True + (n)<sup>§</sup> | 40 | 32 | 38 | 800 | 640 | 768 |
| False - (n)<sup>§</sup> | 10 | 18 | 12 | 200 | 360 | 232 |
| False + (n)<sup>§</sup> | 100 | 0 | 0 | 99 | 0 | 0 |
| True - (n)<sup>§</sup> | 99,850 | 99,950 | 99,950 | 98,901 | 99,000 | 99,000 |
| PPV: true +/all test + (%)<sup>#</sup> | 29% | 100% | 100% | 89% | 100% | 100% |
| Overall accuracy (%)<sup>¤</sup> | 100% | 100% | 100% | 100% | 100% | 100% |
| **Specificity 95% for round 1 tests and 99.9% for rounds 2 and 3** |  |  |  |  |  |  |
| Repeat tests added (n)<sup>¶</sup> | 0 | 5,038 | 5,001 | 0 | 5,750 | 5,105 |
| True + (n)<sup>§</sup> | 40 | 32 | 38 | 800 | 640 | 768 |
| False - (n)<sup>§</sup> | 10 | 18 | 12 | 200 | 360 | 232 |
| False + (n)<sup>§</sup> | 4,998 | 5 | 10 | 4,950 | 5 | 10 |
| True - (n)<sup>§</sup> | 94,953 | 99,945 | 99,940 | 94,050 | 98,995 | 98,990 |
| PPV: true +/all test + (%)<sup>#</sup> | 1% | 86% | 79% | 14% | 99% | 99% |
| Overall accuracy (%)<sup>¤</sup> | 95% | 100% | 100% | 95% | 100% | 100% |
| **Specificity 95% for all tests** |  |  |  |  |  |  |
| Repeat tests added (n)<sup>¶</sup> | 0 | 5,038 | 4,756 | 0 | 5,750 | 4,863 |
| True + (n)<sup>§</sup> | 40 | 32 | 38 | 800 | 640 | 768 |
| False - (n)<sup>§</sup> | 10 | 18 | 12 | 200 | 360 | 232 |
| False + (n)<sup>§</sup> | 4,998 | 250 | 487 | 4950 | 248 | 483 |
| True - (n)<sup>§</sup> | 94,953 | 99,700 | 99,463 | 94,050 | 98,753 | 98,517 |
| PPV: true +/all test + (%)<sup>#</sup> | 1% | 11% | 7% | 14% | 72% | 61% |
| Overall accuracy (%)<sup>¤</sup> | 95% | 100% | 100% | 95% | 99% | 99% |

\* Sensitivity is fixed at 80% for all scenarios. All quoted numbers are expected values, i.e. ignoring the random variation that is inherent to finite samples.
<sup>†</sup>Strategy 1 tests all 100,000 people once
<sup>‡</sup> Strategy 2 tests all people once and then adds a second test (round 2) for those testing positive in their first test (round 1)
<sup>||</sup> Strategy 3 tests all people once, adds a second test (round 2) for those who test positive in round 1 and then a third test (round 3) for those testing negative in round 2
<sup>¶</sup> For strategy 2, number of repeat tests added in round 2; for strategy 3, number of repeat tests added in round 3. A
<sup>§</sup> Rounded to the nearest whole number of people
<sup>#</sup> Rounded to nearest whole %
<sup>¤</sup> Defined as all correct test results (true + and true-) for the relevant testing strategy as a percentage of the final test results from that strategy in all 100,000 people (one result per person only). Rounded to the nearest whole %.

### Interpreting test results for an individual

For the same scenarios, Table 2 shows the results of each testing strategy for an individual, selected at random from the relevant population (and so most likely asymptomatic). Results are shown as the probability (expressed as a percentage) of truly having infection for each set of results based on the individual having had one, two or three tests. The yellow shaded areas of the table show the single or combined test results that would give individuals test positive status based on the three testing strategies. They indicate that, in these low prevalence settings, an individual can only be confident (say, > 90% probability) of test positive status meaning they actually have infection if they have had a confirmatory positive test (i.e. two positive tests) and specificity is extremely high (99.9%) for both tests or - with slightly higher prevalence (1%) – high (95%) for the first test and extremely high (99.9%) for the second. It also shows that if the individual has discrepant results from tests 1 and 2, a third test does not increase the confidence in a positive result, and may substantially reduce it, especially at very low prevalence. For example, if the individual is drawn from a population with 0.05% infection prevalence and test specificities are 95% for the first test and 99.9% for subsequent tests, the probability of being infected given two positive test results is 86.5%; this falls to 56.2% if the first and second tests disagree (positive then negative) and a third is positive (i.e. positive then negative then positive). Finally, when test specificities are high (i.e. 95%) - but not extremely high - throughout, confidence that a positive result implies true infection is poor (probability: 11.4% at best) when population infection prevalence is as low as 0.05% and reasonable but not strong when prevalence is 1%(probability: 72.1% at best).

**Table 2.**
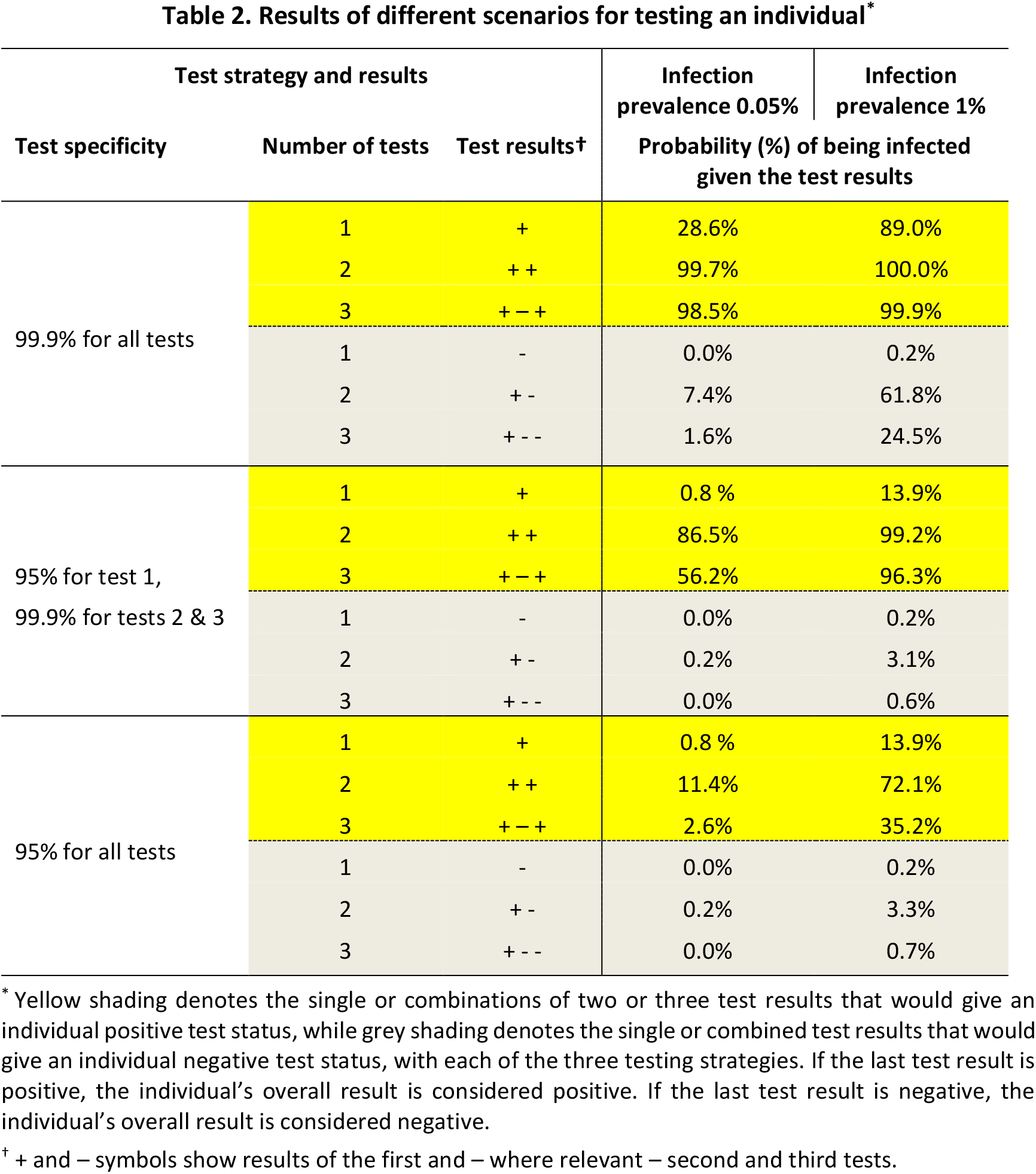
Results of different scenarios for testing an individual^*^

| Test strategy and results |  |  | Infection prevalence 0.05% | Infection prevalence 1% |
| --- | --- | --- | --- | --- |
| Test specificity | Number of tests | Test results<sup>†</sup> | Probability (%) of being infected given the test results |  |
| 99.9% for all tests | 1 | + | 28.6% | 89.0% |
|  | 2 | ++ | 99.7% | 100.0% |
|  | 3 | + - + | 98.5% | 99.9% |
|  | 1 | - | 0.0% | 0.2% |
|  | 2 | + - | 7.4% | 61.8% |
|  | 3 | + - - | 1.6% | 24.5% |
| 95% for test 1,<br>99.9% for tests 2 & 3 | 1 | + | 0.8 % | 13.9% |
|  | 2 | ++ | 86.5% | 99.2% |
|  | 3 | + - + | 56.2% | 96.3% |
|  | 1 | - | 0.0% | 0.2% |
|  | 2 | + - | 0.2% | 3.1% |
|  | 3 | + - - | 0.0% | 0.6% |
| 95% for all tests | 1 | + | 0.8 % | 13.9% |
|  | 2 | ++ | 11.4% | 72.1% |
|  | 3 | + - + | 2.6% | 35.2% |
|  | 1 | - | 0.0% | 0.2% |
|  | 2 | + - | 0.2% | 3.3% |
|  | 3 | + - - | 0.0% | 0.7% |

\* Yellow shading denotes the single or combinations of two or three test results that would give an individual positive test status, while grey shading denotes the single or combined test results that would give an individual negative test status, with each of the three testing strategies. If the last test result is positive, the individual's overall result is considered positive. If the last test result is negative, the individual's overall result is considered negative.
<sup>†</sup> + and – symbols show results of the first and – where relevant – second and third tests.

### Effect of population infection prevalence on the probability that an individual’s positive test result is a true positive

The relationship between population infection prevalence and the probability that an individual’s positive result is a true positive (i.e. the positive predictive value [PPV] for an individual) is shown graphically in Figure 1. Results are shown for a single positive test (Figure 1A) and for two positive tests (Figure 1B), considering different test specificities (or combinations of specificities for two tests). The graphs provide a clear visual illustration of the importance of considering the prevalence of infection in the relevant population or community, the testing strategy (one or two tests required to declare a positive result) and test specificity when interpreting an individual’s test result.

**Figure 1.**
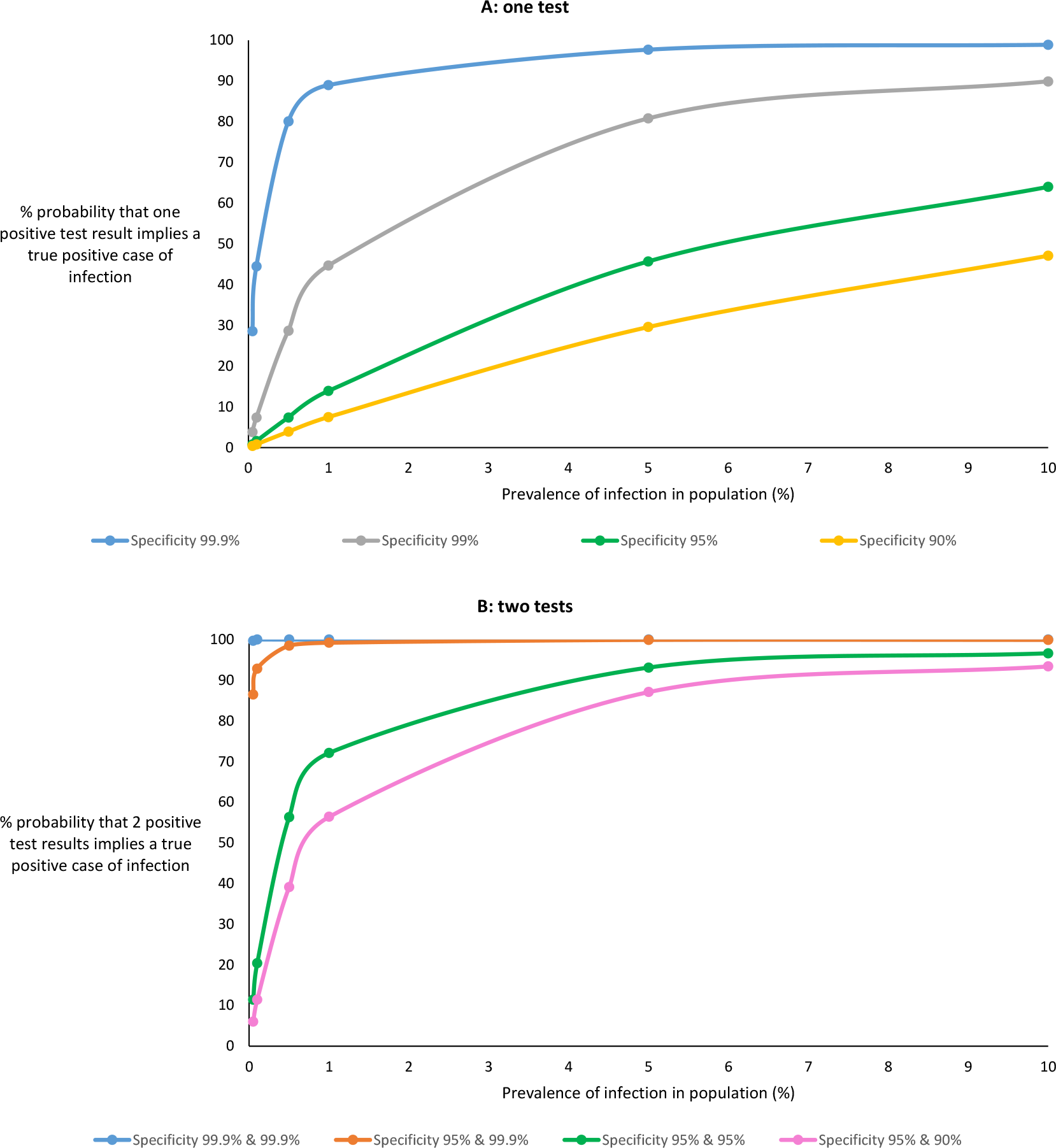
Percentage probability of an individual’s test result being a true positive for different test specificities based on (A) one positive test and (B) two positive tests, at increasing prevalence of current infection in the population from which the individual is drawn

## DISCUSSION

The results of our publicly available interactive tool (https://www.hdruk.ac.uk/projects/false-positives/), which is based on the simple rules of probability, highlight the challenges of dealing with false positive results where large-scale testing of populations is contemplated. We show how in low infection prevalence settings (e.g., unselected asymptomatic school children, asymptomatic people travelling by air or train for work or leisure, or asymptomatic people who have recently attended a pub or restaurant where there was a suspected case of infection), a high percentage of positive results will be false positives, and confidence in the predictive value of an individual’s positive test result will be low, unless both confirmatory testing and extremely high test specificities are in place. This may seem counter-intuitive at first, but a simple thought experiment may help: if you were a random asymptomatic person living in the UK in 2020 and happened to have a test for Ebola virus, would you believe you had Ebola if the test result was positive? Almost certainly not. A key message is to beware of a positive test result when the condition is highly unlikely. It is clearly crucially important to know the likely prevalence of infection before interpreting a population’s or an individual’s test results, or indeed even testing at all.

We also demonstrate that overall accuracy is uninformative and even potentially misleading on the issue of false positives in low prevalence settings, where the vast majority of people being tested are negative. The percentage of all positive results in the tested population that are truly positive is far more informative, given the potential policy implications for the population being tested. Where this percentage is low, most positive results will be false positives and the absolute number of infected cases may be substantially over-estimated. As a result, particular schools, pubs or restaurants may be closed unnecessarily, or lockdown measures affecting towns, cities or regions may be introduced or intensified, again unnecessarily. For individuals with positive results, if the probability that their positive result reflects true infection is low, then mandated quarantine periods for them and their contacts, inability to work or travel, loss of income, and the potential stigma (or worse) of being infected would be unnecessary for most.

A further important result is that there is little – if anything – to gain in low prevalence settings from a strategy that includes a third round of testing aimed at resolving results which are discrepant between the first and second rounds of testing. Indeed, for an individual, the confidence in a positive result may be reduced by such a strategy in comparison with one that includes a second round of confirmatory testing only.

The issue of inaccurate SARS-CoV-2 infection test results has already been considered in the context of testing patients with symptoms, who have a much higher chance of being infected than asymptomatic individuals from low prevalence settings, and where the key problem is that of false negatives rather than false positives.^8^ Others have discussed the problem of high numbers of falsely reassuring false positive results in the context of SARS-CoV-2 *antibody* testing.^9^ A report from the Tony Blair Institute for Global Change, ‘Changing the game on testing’, which calls for population scale testing for SARS-CoV-2 infection, acknowledges that confirmatory testing for those with an initial positive result will be needed, but does not - as we do here - explore the outcomes of different testing scenarios and strategies or emphasise the critical roles of low prevalence and just slightly lower than optimal specificity in driving up the percentage of false positive results, even where confirmatory testing is conducted.^9^

The strengths of our interactive tool include: the lack of any ‘black box’ component – all the probability calculations needed to derive the results are transparently available and the logic is carefully worked through; the flexibility for any user to examine the effects of any combination of the three parameters; the provision of results in formats that are meaningful for testing large groups or populations as well as for interpreting results for individuals; and the ability to assess different repeat testing strategies. However, although simplicity and transparency make our tool attractive, this does result in some limitations. We have assumed that all rounds of testing are independent of each other, whereas in real world situations, there may be characteristics of particular individuals (e.g., swabbing technique for self-administered tests), sample transport systems, or laboratories (e.g. a tendency towards repeated administrative or sample handling errors) that make them inherently more prone to repeatedly inaccurate results. This could be mitigated by ensuring that confirmatory tests are not self-administered, use a different transport mechanism and are conducted in a different laboratory, fully independent of the first, as well as using a different molecular diagnostic test, but this requirement would pose significant logistical challenges. Further, since the proposed testing strategies assume that repeat testing is immediate (and so unaffected by changes in viral detection over the time course of infection), our tool is only applicable for tests with very fast turnaround of results. These are now becoming available,^10^ but a detailed understanding of their performance, including sensitivity and specificity, will be required to assess their potential use in widespread testing among asymptomatic people.

In conclusion, we agree with others that population-wide testing may have an important role to play in determining local, regional and national policy for the population and in providing guidance (or even mandatory rules) for individuals. However, the crucial importance both of extremely high test specificity and of confirmatory testing must be fully appreciated and incorporated into policy decisions if we are to avoid multiple unnecessary restrictions on individuals and on populations.

## Data Availability

All relevant data pertinent to the manuscript is available within the manuscript or at the links provided.

https://www.hdruk.ac.uk/projects/false-positives/

